# Real-time evaluation and adaptation to facilitate rapid recruitment in a large cohort

**DOI:** 10.1101/2023.01.30.23285102

**Authors:** Ashley Honushefsky, Eric S. Wagner, Kathleen Sheridan, Kathleen M. Spickard, William R. LeMasters, Carroll N. Walter, Taryn Beaver, Alanna Kulchak Rahm, Adam H. Buchanan

## Abstract

**Objectives:** Recruiting large, diverse cohorts efficiently can speed the translation of findings into care across a range of scientific disciplines and medical specialties. Yet, efficient recruitment can be hampered by factors such as financial barriers, logistical concerns, and lack of resources for patients and clinicians. Here we use a case study of a large, prospective trial of a multi-cancer early detection test to describe how the study team tracked enrollment, protocol fidelity, and participant experience and iteratively adapted procedures.

**Methods:** While conducting a large, prospective trial of a multi-cancer early detection blood test at Geisinger, an integrated health system in central Pennsylvania, we monitored recruitment progress, fidelity to protocol procedures, and participants’ satisfaction. Tracking mechanisms such as paper records, electronic health records, research databases, dashboards, and electronic files were utilized to measure each outcome. We then reviewed study procedures and timelines to list the implementation strategies that were used to address barriers to recruitment, protocol fidelity and participant satisfaction.

**Results:** We enrolled 10,006 women ages 65-75 over 22 months. Adaptations to recruitment and enrollment methods that contributed to achieving the enrollment goal included adopting group consenting, improving visit convenience, increasing electronic capture and tracking of data and source documents, staffing optimization via leveraging resources external to the study team when appropriate, and integrating the disclosure of study results into routine clinical care without adding unfunded work for physicians. We maintained high protocol fidelity and positive participant experience as exhibited by a very low protocol deviation rate and a low number of participant complaints.

**Conclusion:** Recruiting rapidly for large studies – and thereby facilitating clinical translation – requires a nimble, creative approach that marshals available resources and changes course according to data. Planning a rigorous assessment of a study’s implementation outcomes prior to study recruitment can further ground study adaptations and facilitate translation into practice. This can be accomplished by proactively and continuously assessing and revising implementation strategies.

**Strengths and limitations of this study:**

- Synthesis and tracking of various data
- Real-time identification of necessary adaptations
- Mapping of adaptations to problems and consequences
- Analysis of results post-hoc
- Inability to analyze the value or impact of a single adaptation

## INTRODUCTION

Recruiting large, diverse cohorts in a relatively short period of time can speed the translation of findings into care across a range of scientific disciplines and medical specialties. For example, SARS-CoV-2 vaccine trials demonstrated the potential for rapid public health impact when trials reach enrollment goals quickly and effectively.^1-2^ Rapid enrollment of large cohorts is also important in studies with regulatory implications that are not as imminent as a vaccine for a global pandemic but require substantial cohorts to achieve the power necessary to compare clinical outcomes, and in studies that seek to elucidate associations between genes and disease.^3-4^ Potential benefits of rapidly enrolling large cohorts include a reduction in study costs for the sponsor and a shortened timeline for an FDA-approved product to reach the market.^5^

Yet, factors such as financial barriers, logistical concerns, and lack of resources for patients and clinicians to support clinical trial enrollment and retention can prevent efficient enrollment, particularly when studies do not have the resources or organization to overcome these barriers.^6^ Further, few studies have the resources that have been available for SARs-CoV-2 vaccine trials or large industry- or government-sponsored initiatives, meaning that most studies must accomplish recruitment goals using the more limited resources on hand. Establishing procedures prior to trial start-up can maximize the success of trial implementation.^7^ However, challenges arising throughout implementation may still threaten recruitment, retention, adherence to the treatments or interventions being studied, and data collection, negatively impacting fulfillment of study aims. Therefore, the ability to identify and address such challenges in real-time is critical.

The DETECT-A study is an interventional, single-group study in which all participants had a liquid biopsy venipuncture for a multi-cancer early detection test (called CancerSEEK) and proceeded to whole body PET-CT scan for tumor localization if CancerSEEK was abnormal.^8^ Enrollment and recruitment for DETECT-A was conducted at Geisinger, a single, partially integrated healthcare system spanning 45 counties in Pennsylvania, 35 of which are designated as rural. Geisinger was chosen as the recruitment site due to several features that were anticipated to facilitate recruitment. By virtue of its large, stable patient population and 25-year use of an electronic health record that can be queried for potentially eligible individuals,^9^ investigators hoped to enroll 10,000 participants in 18 months. This paper describes how the study team identified challenges during recruitment and enrollment and implemented solutions that allowed the study to reach the enrollment goal while maintaining protocol fidelity and facilitating a positive participant experience.

## METHODS

### DETECT-A Study Overview

DETECT-A evaluated the feasibility and safety of incorporating a multi-cancer early detection blood test into routine clinical care.^8^ To analyze these outcomes, the study was designed to enroll 10,000 women ages 65-75 who had no prior history of cancer. Recruitment began in August 2017 and enrollment was anticipated to be completed in 18 months. Study procedures, which are described in detail elsewhere,^8^ are summarized in Figure 1. Recruitment efforts, baseline enrollment visits, and follow-up activities overlapped chronologically, making it important to quickly assess and adapt strategies to ensure that recruitment goals, protocol fidelity, and patient satisfaction were simultaneously achieved (Figure 2). Here we report the initial procedures for achieving the recruitment goal and methods for tracking study processes. Adaptations based on the results of process tracking are summarized in Results.

**Figure 1.**
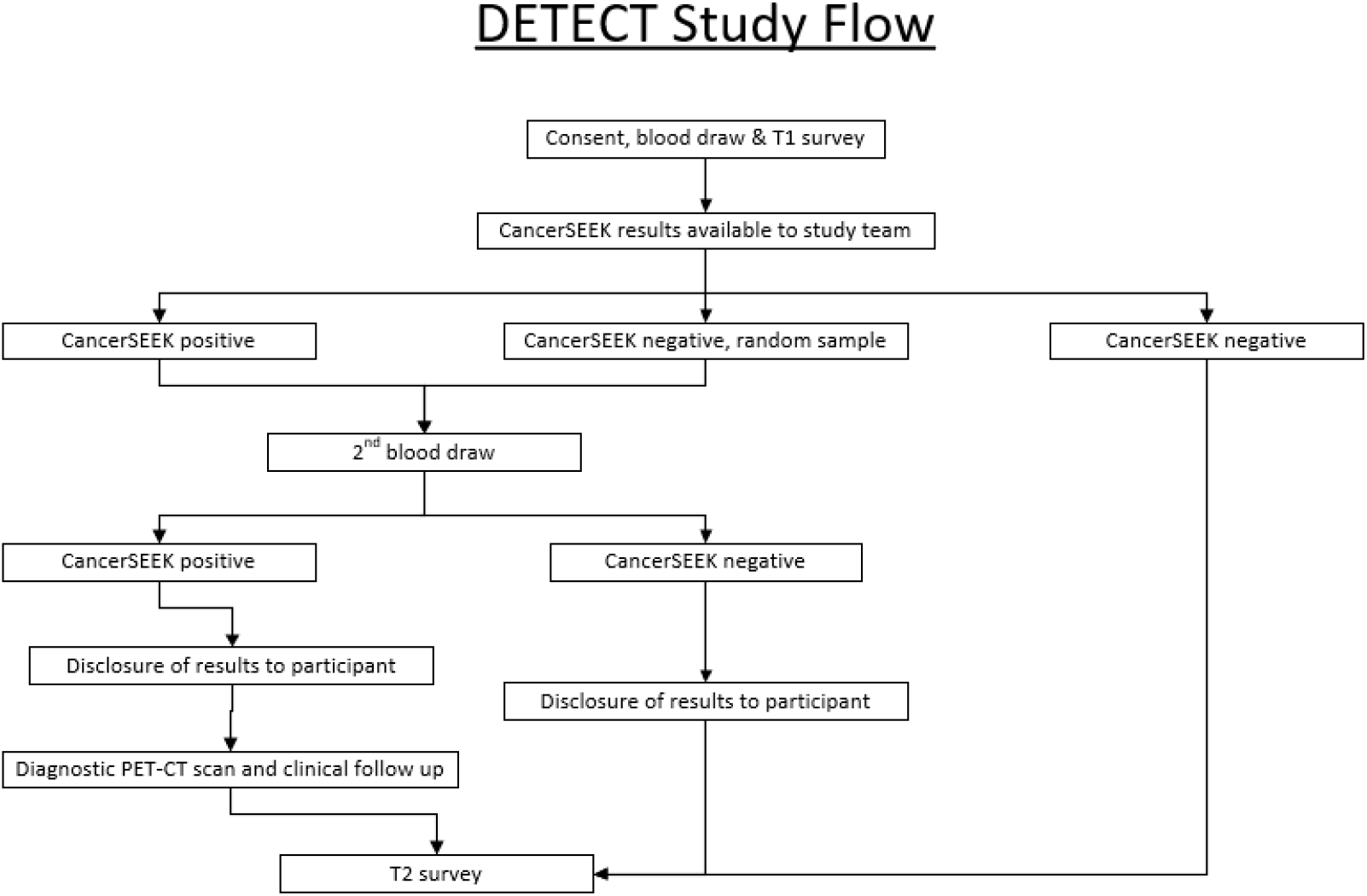
DETECT-A Study Design Summary.

**Figure 2.**
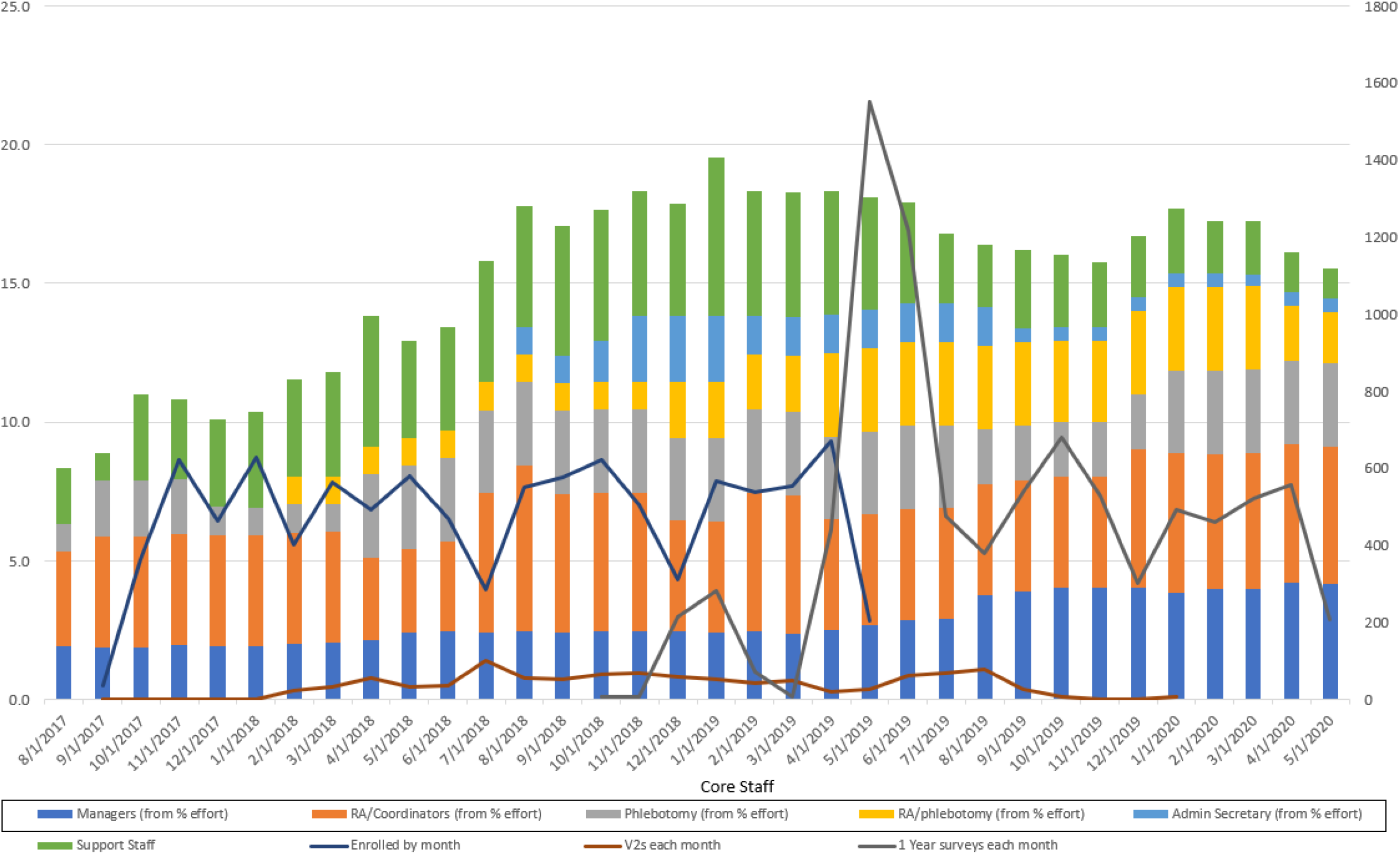
Enrollment and 1-year Follow-up Timeline Showing Staffing Needs.

### Initial Plan

The initial study team comprised two project managers, four research assistants, and one research phlebotomist, all of whom were fully funded by the study. Enrollment commenced at two locations where study staff had permanent office and/or clinic space. Initial recruitment methods included targeted mailings, flyers, and referrals from study staff and participants. Targeted mailings were sent in 101 batches (of fewer than 500 letters) to potentially eligible individuals who were identified by querying Geisinger’s electronic health record (EHR). Letters included return postcards for recipients to express their interest or disinterest in participating.

### Measures of Implementation

Tracking and monitoring techniques to assess the recruitment progress, fidelity to protocol procedures, and participants’ satisfaction are summarized in Tables 1 and 2, and below.

**Table 1.** Task Prioritization by Team.

|  | VISIT TEAM | OFFICE TEAM |
| --- | --- | --- |
| HIGH PRIORITY, TIME SENSITIVE | Staffing Sign-up Sheet<br>Visit Prep Checklist<br>Participant Check-in Process<br>Participant Screening (Inclusion/Exclusion Form)<br>Informed Consenting Process<br>Specimen Collection and Documentation (Phlebotomy Form)<br>Laboratory Manifest Completion<br>Baseline Survey Administration<br>Giftcard Distribution<br>Specimen Shipment | Managing incoming opt-in post cards<br>Retrieving Voicemails (Communicating with Visit Team)<br>Distribution of Outgoing Calls<br>Incoming Calls (Communicating with Visit Team)<br>Generating Appointment Sheets<br>Reminder Calls for Upcoming Appointments<br>Finalizing and Printing Appointment Sheets<br>Laboratory Manifest Auditing (Real time)<br>Documenting Visits in EHR (Real time)<br>Sending Laboratory Manifest |
| LOWER PRIORITY, TIME FLEXIBLE | Baseline Survey Entry into REDCap<br>Phlebotomy Form Entry into REDCap<br>Visit Paperwork Auditing | CancerSEEK Results Entry into REDCap<br>Positive CancerSEEK Results Scanning into EHR<br>ICF Scanning into EHR<br>1-Year Follow-Up Survey Administration<br>Survey Entry into REDCap<br>Data Entry (CRC Follow-Up and Data Pulls)<br>Giftcard Documentation<br>Inventory and Ordering of Supplies<br>Generating Barcodes for Specimen and Giftcard Tracking<br>Creating Study Metrics Using Power BI Dashboards<br>Financial Tracking and Reconciliation<br>Filing Paper Source Documents<br>Tracking and Addressing Participant Complaints<br>Tracking Metrics for IRB Continuing Review Submissions |

**Table 2.** Data Collection and Tracking Mechanisms.

| **Task** | **Paper** | **EHR** | **REDCap** | **Electronic File** |
| --- | --- | --- | --- | --- |
| Opt-in Postcards | S |  | C |  |
| Staffing Sign-up Sheet |  |  |  | S |
| Visit Prep Checklist | S |  |  |  |
| Check-in Documentation | S | C | C |  |
| Screening (Inclusion/Exclusion) | S |  |  | C |
| Consent Documentation | S | C | C | C |
| Specimen Manifest* |  |  |  | S |
| Phlebotomy Documentation | S |  | C |  |
| Baseline Survey | S |  | C | C |
| Giftcard Distribution | S |  |  | C |
| Incoming and Outgoing Call Documentation |  |  | S | C |
| Appointment Tracker | C | C | C | S |
| CancerSEEK Results |  | C | C | S |
| Annual Follow-up Surveys | S |  | S | C |
| CRC Data |  |  | C | S |
| Inventory and Ordering | C |  |  | C |
| Printing Barcodes for Specimen and Giftcard Tracking | C |  |  | S |
| Creating Study Metrics Using Power BI Dashboards |  |  |  | S |
| Tracking and Addressing Participant Complaints |  |  | S | C |
| Tracking Metrics for IRB Continuing Review Submissions |  |  |  | S |
| *S = source documentation in which data collected was first recorded*<br>*C = copy of data documentation that was later entered into an electronic data capture system* |  |  |  |  |

#### Recruitment procedures

Research Electronic Data Capture (REDCap)^10^ databases were used to manage and monitor participant interest and progression throughout the study and provide weekly screening and enrollment metrics to the study collaborators (Figure 3). The study team used REDCap data to create Power BI dashboards to routinely assess important metrics such as the number of unfilled informed consent discussion appointment slots and recruitment and follow-up call assignments. If appointment slots were not being filled adequately, recruitment strategies and staff priorities were adjusted as needed (see Results). When potential participants were not interested in attending an enrollment event at currently available locations, we tracked their preferred location in REDCap and considered adding these locations as enrollment sites.

**Figure 3.**
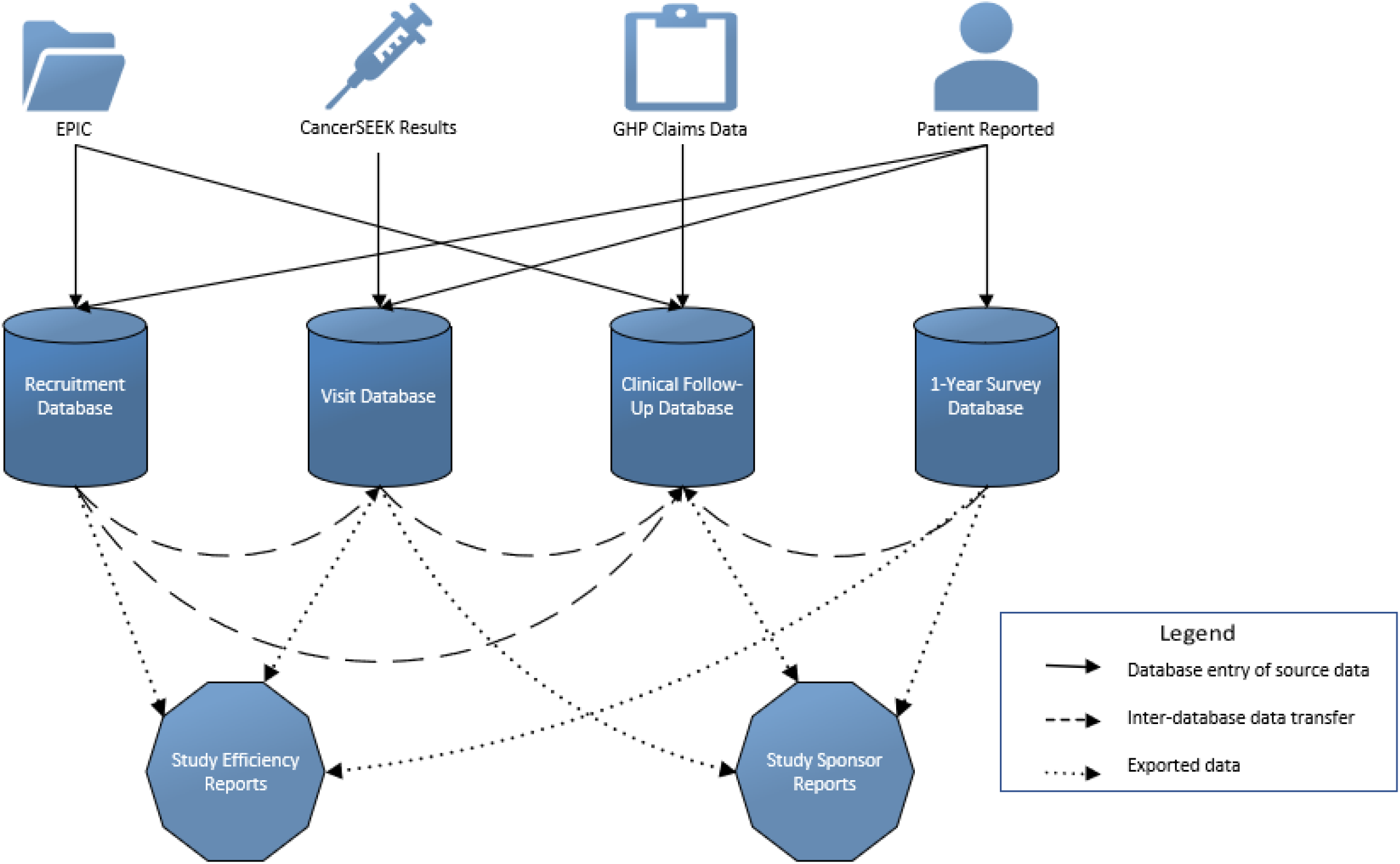
Outline of Databases, Dashboards and Reports.

#### Protocol fidelity procedures

Tracking and monitoring of protocol activities that occurred throughout the study were complex, requiring flexibility and coordination of study staff. Staff members were divided into two ‘teams’ that had different priorities and tasks (Table 1). Each team was responsible for monitoring their daily tasks to help with problem-solving in real time. Study activities were collected and tracked on paper, in the participants’ EHR, in the study’s REDCap database, and in electronic files (Table 2). In addition to tracking and monitoring, the team was trained to identify potential issues proactively so that we could adapt to situations as needed. Protocol deviations and PHI breaches were handled immediately, documented in electronic spreadsheets and REDCap databases, and reported to the IRB at continuing review.

#### Participant Experience Procedures

Throughout the study, participants were encouraged to contact the study team, a study genetic counselor, or Geisinger’s IRB with any problems, questions, or concerns. Complaints about the sites where the visits were held were reported to a study coordinator.

## RESULTS

The DETECT-A study enrolled 10,006 participants over the course of 22 months, which was four months longer than our anticipated timeline. Table 3 shows a monthly breakdown of adaptations to the recruitment methods and locations used to achieve the enrollment goal. Adaptations to our recruitment, protocol, and participant experience processes are summarized in the tables below (Tables 3 and 4).

**Table 3.** DETECT-A Recruitment Timeline

| Month of Recruitment | Recruitment Methods |  |  |  |  |  |  | Enrollment Metrics |  |  |  |
| --- | --- | --- | --- | --- | --- | --- | --- | --- | --- | --- | --- |
|  | Mass Mailings (M)<sup>1</sup> | Referrals (R)<sup>2</sup> | Flyers (F)<sup>3</sup> | Social Media (S) | News Media (NM) | Newsletters (N) | Community Outreach Events (C) | Locations Used | Locations Available (cumulative) | Enrolled per Month | Enrolled (cumulative) |
| M1 Aug | M<sup>1</sup> | R<sup>1</sup> | F<sup>1</sup> |  |  |  |  | 0 | 0 | 0 | 0 |
| M2 Sep | M<sup>1</sup> | R<sup>1</sup> | F<sup>1</sup> |  |  |  | C | 2 | 2 | 36 | 36 |
| M3 Oct | M<sup>1</sup> | R<sup>1</sup> | F<sup>1</sup> | S | NM |  |  | 4 | 4 | 364 | 400 |
| M4 Nov | M<sup>1</sup> | R<sup>1</sup> | F<sup>1</sup> | S | NM |  |  | 3 | 4 | 621 | 1021 |
| M5 Dec | M<sup>1</sup> | R<sup>1</sup> | F<sup>1</sup> | S |  |  | C | 4 | 4 | 464 | 1485 |
| M6 Jan | M<sup>1</sup> | R<sup>1</sup> | F<sup>1</sup> |  |  |  |  | 4 | 5 | 629 | 2114 |
| M7 Feb |  | R<sup>1</sup> | F<sup>1</sup> | S |  |  |  | 5 | 5 | 403 | 2517 |
| M8 Mar |  | R<sup>1</sup> | F<sup>2</sup> |  |  | N |  | 7 | 7 | 564 | 3081 |
| M9 Apr |  | R<sup>1</sup> | F<sup>2</sup> |  | NM |  |  | 8 | 8 | 491 | 3572 |
| M10 May |  | R<sup>1</sup> | F<sup>2</sup> | S |  |  |  | 9 | 14 | 581 | 4153 |
| M11 Jun |  | R<sup>1</sup> | F<sup>2</sup> | S |  |  |  | 14 | 18 | 470 | 4623 |
| M12 Jul |  | R<sup>1</sup> | F<sup>2</sup> |  |  |  |  | 10 | 18 | 284 | 4907 |
| M13 Aug |  | R<sup>1</sup> | F<sup>2</sup> |  | NM | N |  | 14 | 18 | 552 | 5459 |
| M14 Sep | M<sup>2</sup> | R<sup>1</sup> | F<sup>2</sup> | S |  |  | C | 15 | 18 | 577 | 6036 |
| M15 Oct | M<sup>2</sup> | R<sup>1</sup> | F<sup>2</sup> | S |  | N | C | 16 | 20 | 666 | 6702 |
| M16 Nov | M<sup>3</sup> | R<sup>1</sup> | F<sup>2</sup> | S |  | N | C | 16 | 21 | 462 | 7164 |
| M17 Dec | M<sup>3</sup> | R<sup>2</sup> | F<sup>2</sup> | S |  | N | C | 15 | 21 | 310 | 7474 |
| M18 Jan | M<sup>3</sup> | R<sup>2</sup> | F<sup>2</sup> | S | NM | N |  | 15 | 22 | 566 | 8040 |
| M19 Feb | M<sup>3</sup> | R<sup>2</sup> | F<sup>2</sup> |  |  | N |  | 15 | 22 | 537 | 8577 |
| M20 Mar | M<sup>3</sup> | R<sup>2</sup> | F<sup>2</sup> |  |  | N |  | 16 | 22 | 555 | 9132 |
| M21 Apr |  | R<sup>2</sup> | F<sup>2</sup> |  |  | N |  | 16 | 22 | 670 | 9802 |
| M22 May |  | R<sup>2</sup> | F<sup>2</sup> |  |  | N |  | 14 | 22 | 204 | 10006 |

<sup>1</sup> Initial mass mailings (M<sup>1</sup>) included an invitation letter and a return postcard. At month 14, the return postcard was replaced by an information card (M<sup>2</sup>) that featured a photo of study participants. At month 16, the details from both the information card and invitation letter were incorporated into a single invitation card (M<sup>3</sup>).
<sup>2</sup> Initial referrals (R<sup>1</sup>) were accepted from physicians, research staff, and participants. At month 17, we added a Refer-A-Friend program (R<sup>2</sup>) until enrollment ended.
<sup>3</sup> Initial flyers (F<sup>1</sup>) included only text and were printed by study staff. At month 8, Geisinger's Marketing and Communications department designed flyers (F<sup>2</sup>) with stock photos and tear-off contact information.

**Table 4.**
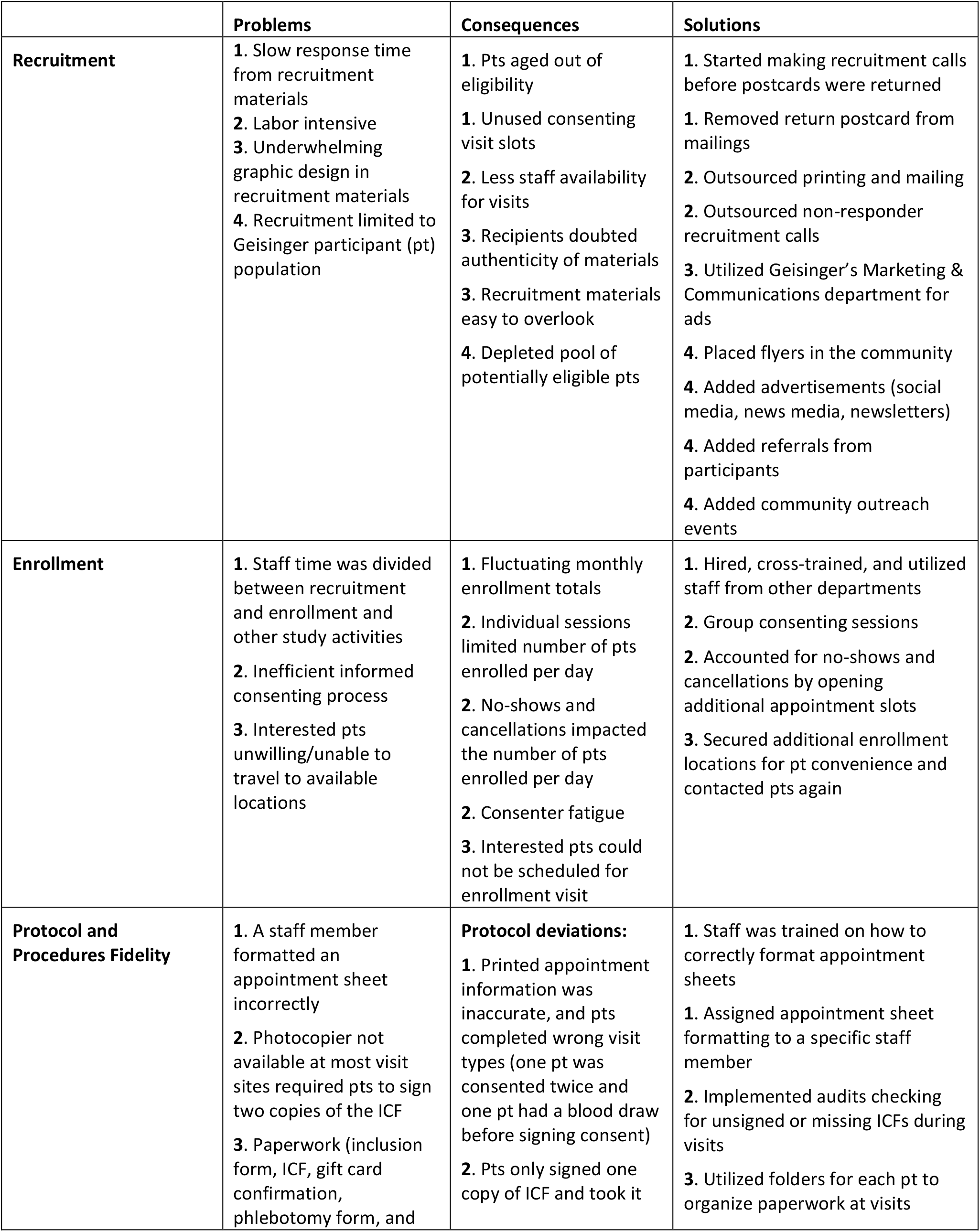
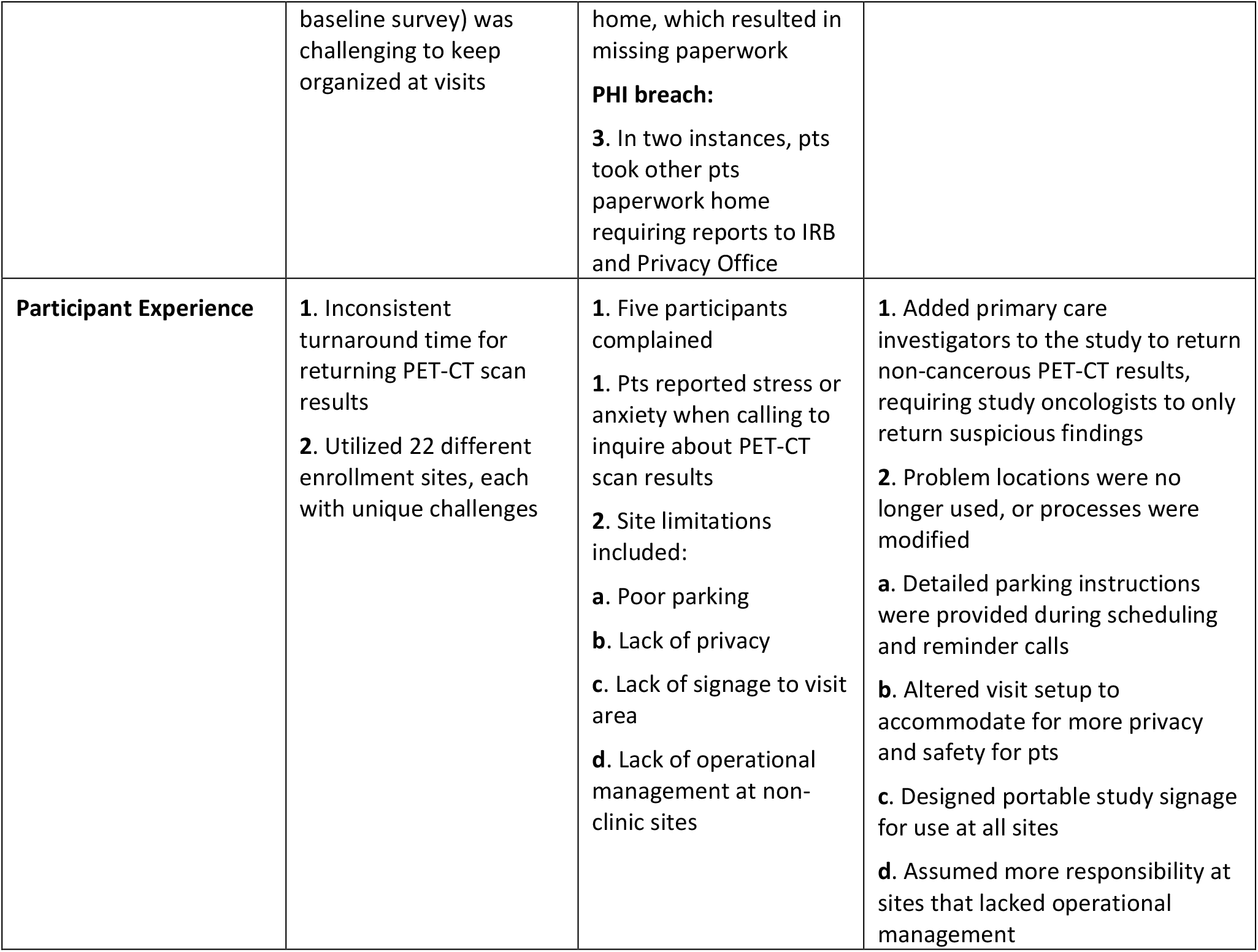
Rapid Cycle Evaluation of Study Implementation.

### Recruitment procedures – Problems and Consequences

The initial recruitment methods used in the first two months (Table 3) presented several problems that impacted potential to reach the recruitment target. The variability in the return time of postcards indicating respondent interest led to potential participants no longer meeting eligibility criteria, which impacted our ability to fill appointment times at study visits. It was labor intensive for the staff to manage postcards, which limited their availability for visits. The graphic design of the initial recruitment materials (i.e., invitation letter and return postcard) were bland and did not elicit interest from recipients who doubted their authenticity. Mailings were limited to Geisinger patients, which resulted in a smaller pool of potentially eligible participants early in the study. Together, mailings and flyers alone were insufficient in both generating enough interested participants and in generating them quickly enough to meet our weekly enrollment targets.

### Recruitment procedures - Solutions

To reduce response turnaround time from the initial recruitment methods, modifications were made to the targeted mailing process and to the flyers. Postcards were eliminated to make the recruitment process more time efficient. Geisinger’s Marketing & Communications department created recruitment content with more engaging graphic design. We also supplemented our recruitment efforts with methods designed to reach non-Geisinger patients, including advertisements (social media, news media, newsletters, flyers), referrals from participants, and community outreach (Table 3). Both targeted and broad recruitment methods were used in tandem as necessary to meet weekly recruitment targets (Figure 4).

**Figure 4.**
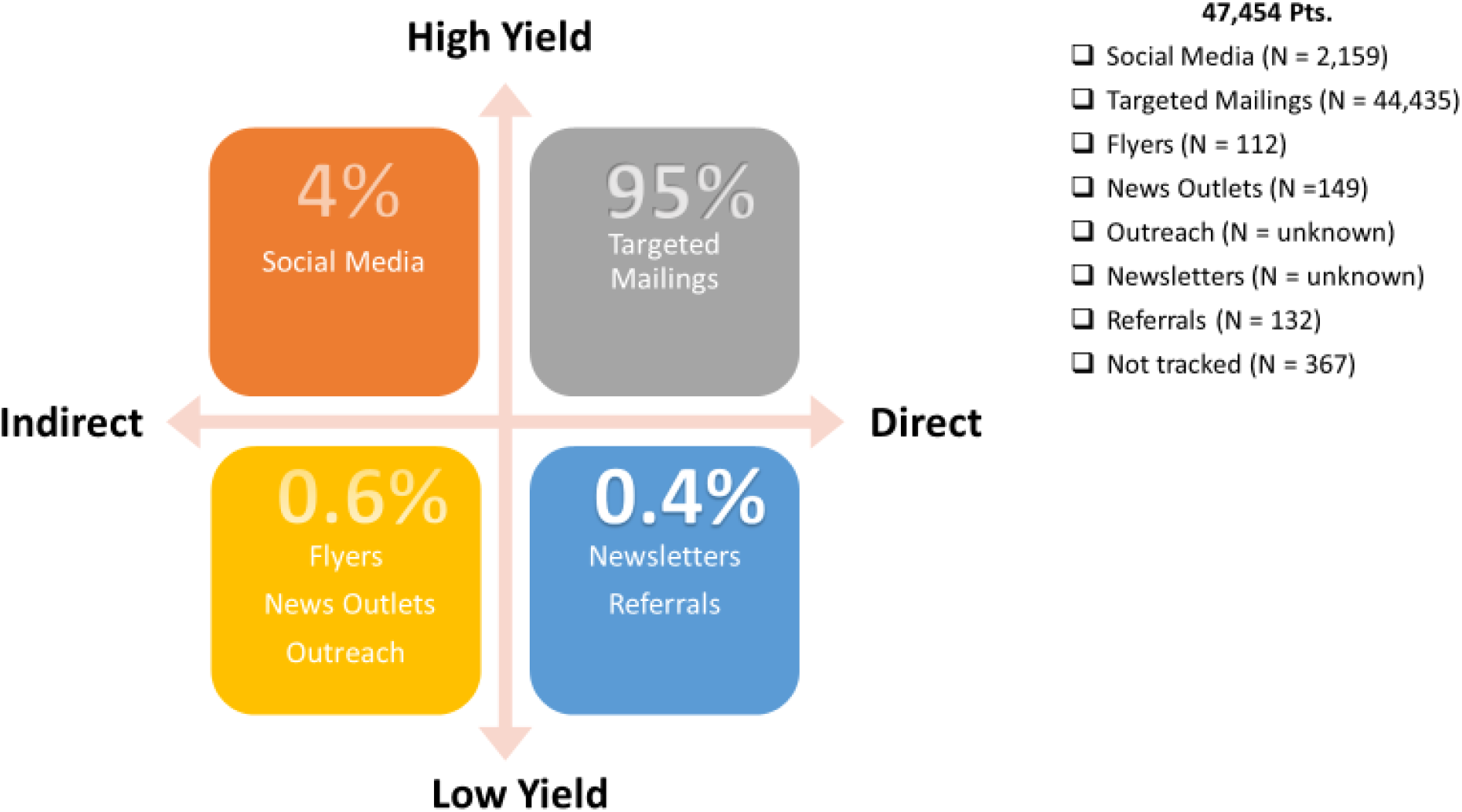
Percent Yield.

**Figure 5.** Percentages shown above represent the number of participants who enrolled in the study by recruitment method category. Targeted recruitment methods focused on individuals who were more likely to meet inclusion/exclusion criteria. Broad methods reached a larger population that resulted in a lower percentage of eligible individuals. Mass mailings and social media advertisements resulted in the highest yield of interested and eligible individuals.

Facebook advertisements were created using demographic and geographic parameters. Readers who clicked the ad were routed to the DETECT-A website, where they could complete an eligibility questionnaire or find contact information for the study team. The study team reviewed the eligibility of each submission, including EHR review for Geisinger patients. An email was sent to those who were not eligible. Those who were potentially eligible were called and inclusion/exclusion criteria were reassessed. Social media recruitment, which had a link to opt-in electronically that directly alerted the study staff, had the quickest response time compared to other recruitment methods.

Study staff participated in multiple community outreach events where the target population was represented, such as local community fairs, health and wellness fairs, and community luncheons for older individuals. Stories about the DETECT-A study and its investigators were featured in local newspapers, on Geisinger-sponsored health segments, and on local news programs. Upcoming enrollment events were highlighted in standing communications from Geisinger to members of the Geisinger Health Plan (GHP)^11^, Silver Circle^12^, and other large Geisinger research projects like the MyCode Community Health Initiative.^13^

Flyers were distributed in the community and participants were encouraged to share study flyers with other potential participants. We also ran a “Refer-a-Friend” mailing campaign encouraging participants to refer eligible friends and family. When a new potential participant contacted the study team, the participant who referred them was entered into a gift card drawing, which occurred at the close of enrollment.

### Enrollment procedures - Problems and Consequences

Enrolling the eligible and interested individuals presented additional challenges beyond those encountered during recruitment. Interested individuals were sometimes unwilling or unable to travel to available enrollment locations to provide informed consent, negatively impacting enrollment numbers. Individual informed consent sessions resulted in consenter fatigue, a lower daily capacity for enrollment, and a loss in productivity when potential participants canceled or did not show for scheduled appointments. Recruitment efforts and enrollment events competed for staff time, which was reflected in fluctuating enrollment numbers from month to month. Months with heavy enrollment meant staff were largely deployed to visits, which were then followed by months with unfilled appointment slots as staff had less time to devote to recruitment calls (Figure 2).

### Enrollment procedures - Solutions

We tailored teams to meet recruitment, enrollment, and other study activity demands (Table 1). Visit logistics were designed to make the Visit Team mobile. Over the 22 months of recruitment, locations were expanded to a total of 22 sites across the Geisinger service area based on feedback from recruitment calls (Table 3). The spaces to which we were given access consisted of exam rooms, conference rooms, multipurpose rooms, employee breakrooms, community rooms, the Geisinger Commonwealth School of Medicine, an athletic facility, and an indoor courtyard. Staff traveled in study vehicles to the enrollment locations with tables, chairs, lab kits, phlebotomy supplies (including snacks and water for participants post blood draw), signage, and paperwork.

Group consenting sessions with two to 15 potential participants were instituted, allowing study personnel to consent two to five times as many participants per day (up to 103 participants) with adequate rest breaks between sessions. Staff were available if a participant preferred an individual consenting session. Group sizes were predetermined based on the layout of each site and were scheduled accordingly. The flexibility in group consenting sessions allowed us to overbook the schedule to account for no-shows and cancelations.

We cross-trained fully funded staff and employed research support department staff to assist with recruitment, conducting informed consent, and blood draws, which freed up fully funded staff for office tasks (Figure 2). We utilized Geisinger’s Survey Research Core to assist with recruitment calls, scheduling, and survey administration, allowing us to expand these activities to nights and weekends. We were able to leverage additional system-wide internal resources, such as having Research Assistants train in Geisinger’s School of Phlebotomy to perform venipunctures, which expanded our team of research phlebotomists and improved visit efficiency.

In total, 111 employees billed their time and effort to the study during the 22 months of recruitment and the first 12-months of follow-up, which translated to an average of 15.29 full-time employees. This included core study staff, genetic counselors, physicians, research call center and other support staff. These diverse staffing resources allowed for division of labor by area of expertise. Additionally, our core staff was cross trained for both office and visit tasks (Table 1), which gave us the flexibility to adapt procedures as the study demanded.

### Protocol fidelity procedures - Problems and Consequences

We had four types of protocol deviations: using outdated informed consent (ICF) versions, enrolling more participants than were IRB-approved, misplacing original ICFs, and signing an ICF after completing venipuncture. There were two types of protected health information (PHI) breaches reported to the privacy office, including a mix-up of paperwork at the visits, and mailing paperwork to the wrong participant. There were two issues of non-compliance for unauthorized HIPAA use for internally providing a patient list that included PHI to a statistician who was not yet added to IRB study application and accidental sharing of unapproved PHI on an internal and private group communication board. Table 4 details the problems that resulted in adaptations to our processes. The problems and consequences were largely related to the use of paper source documents, which were inefficient, time-consuming, and vulnerable to errors.

### Protocol fidelity procedures - Solutions

#### Protocol Deviations

To avoid future protocol deviations, it was necessary to periodically retrain staff on certain processes. For complex and technical tasks, we designated a point person to have primary responsibility to decrease protocol errors. For example, an assigned staff member exported reports to generate appointment sheets, which led to better organization and easier identification of participants’ visit types at each visit. Multiple staff were retrained as backups and were used when needed on that assignment. Identification of visit types were key because participants were at different stages of the study. For instance, a baseline visit included the informed consent process, questionnaire, and blood draw versus follow up blood draw or redraw appointments for those participants who were already consented into the study.

We implemented checks throughout the visit to ensure the ICF was signed and filed appropriately, which enabled us to identify missing signatures or forms and obtain them before the participant left the visit.

#### PHI Breaches

To facilitate fidelity to the protocol procedures, each participant was given a folder at the start of the visit containing the inclusion/exclusion verification sheet, gift card confirmation, and phlebotomy form, as well as the ICF and baseline survey. The participant kept the folder with them throughout the visit. At the conclusion of their visit, study staff retrieved original paperwork from the folder and returned copies of the ICF and gift card confirmation form to the participants.

### Participant experience procedures - Problems and Consequences

Four of the 22 enrollment sites presented challenges that were identified by participants using a patient-satisfaction survey. Dissatisfaction included comments regarding limited or inconvenient parking, privacy concerns, lack of signage directing potential participants to visit areas, and limited facility oversight at non-clinic sites. Of clinical importance, five participants who underwent a PET-CT scan reported anxiety due to prolonged wait times between imaging and result disclosure. Also, there were complaints related to study design. For example, three participants that had a negative CancerSEEK test were upset they did not hear from the team again until it was time to complete their 1-year follow up survey. We also had three participants who were upset that they were diagnosed with cancer despite their CancerSEEK test being negative.

### Participant experience procedures - Solutions

We discontinued use of enrollment sites that prompted multiple participant complaints. To improve participant experience at remaining sites, we altered visit set-ups, provided more detailed parking instructions, and displayed portable study signage. It was necessary for the study team to take more responsibility at sites that lacked operational management.

We increased the number of clinician investigators on the study to assist with the timely return of PET-CT scan results. Study staff triaged the findings to the appropriate study physician. Specifically, primary care physicians on the study were enlisted to return imaging findings that were not concerning for cancer. This allowed study oncologists to focus on only returning the findings suggestive of cancer. In all instances, participants’ primary care physicians were also notified of the results and recommended next steps via EHR message, fax, or phone call. We did not address study design concerns participants raised but we did offer counseling visits for those who were upset with results or new diagnoses.

## DISCUSSION

The effective and efficient recruitment of large numbers of individuals over short time periods is critical to translating research findings into practice.^3-5^ The lessons learned from research studies like DETECT-A can inform future large recruitment efforts and foreshadow important clinician or individual patient-level implementation obstacles. Our findings regarding successful recruitment strategies and the importance of data-driven adaptations to these strategies are consistent with recent recruitment literature.^14-17^ In particular, iterative adaptation to a priori recruitment and enrollment strategies based on timely evaluation of available data was key to our study’s ability to meet enrollment targets.^17^ Using existing data capture systems like REDCap^10^ can streamline the ease with which data from these recruitment strategies can be analyzed.^18^ We also realized the importance of a patient-centered approach to conducting the trial, which allowed us to respond to participant-reported barriers to recruitment and enrollment. Consistent with the work of Masese et al. among people of African descent with sickle cell disease, using a variety of recruitment strategies is likely to be necessary for effective recruitment, particularly when attempting to overcome recruitment barriers among populations underrepresented in research.^16^ Using multiple strategies (e.g., offering informed consent processes by multiple modalities such as telephone, chatbot, telehealth, and in-person) also offers opportunities to be as inclusive as possible and limit selection bias.

The most impactful adaptations to recruitment strategies were related to group consenting, staffing, and participant experience. These solutions may be effective within other study designs and organizational contexts. Initially, the study used individual consenting sessions, as is typical in many studies. It was quickly apparent from enrollment figures that this approach was inefficient and recruitment goals would not be reached. Shifting to a group consenting format resulted in efficiencies that facilitated the ability to reach goals on target without negatively impacting fidelity to study protocol. Anecdotally, we observed that group consenting enhanced conversational dynamics and allowed for deeper and more meaningful discussion of the informed consent form, an experience consistent with the impact of group dynamics in clinical settings.^19-22^ Geisinger’s service area is predominantly rural and covers a large geographic area, which sometimes required participants to travel long distances to reach an enrollment location. Our expanding recruitment efforts and study activities required a rapid increase in study staff, and the increased staff size allowed us to expand to many of the requested locations that were more convenient for participants. Some of these locations were in less populous areas, so by scheduling at more than one site per day we were able to use staff time efficiently while meeting daily enrollment targets. The study leased two vehicles for staff travel to cover the additional visit sites. Given that recruitment was completed prior to the COVID-19 pandemic, further strategies (e.g., telehealth consenting) are now useful options.

As we adapted recruitment strategies during the study, we realized, as others have, the importance of considering implementation strategies and associated outcomes while planning a study.^14^ In fact, though we did not intend to do so from the outset, we used several of the implementation strategies named by Powell et al., including changing service sites, developing and implementing tools for quality monitoring, and promoting adaptability.^23^ Using an implementation science framework and associated list of implementation strategies from the beginning of the study would have improved our ability to identify and evaluate the impact of adaptations to recruitment strategies. It might have also allowed us to better address another study limitation - the inability to analyze the value or impact of one single process change, since many adaptations were implemented simultaneously due to aggressive study timelines that required us to address multiple issues at once. Despite these limitations, we believe that the post-hoc analysis of implementation outcomes described in this manuscript is valuable for informing future measurements and tracking adaptations.

Based on these experiences in the DETECT-A study, we recommend instituting the following from the outset of a clinical research study: capturing and tracking data and source documents electronically as much as possible, leveraging resources external to the study team when appropriate, and, for interventional trials, integrating the disclosure of study results easily and seamlessly into routine clinical care without adding unfunded work for physicians.^15^ Extensive electronic tracking of study processes allows the team to identify ineffective processes, adapt quickly based on data, and avoid protocol deviations. Another important lesson was to not undertake tasks for which other groups are experts, such as utilizing Marketing and Communications and the Digital Print and Mail Center in the design of posters and other recruitment materials.

Recruiting rapidly for large studies – and thereby facilitating clinical translation – requires a nimble, creative approach that marshals available resources and changes course according to data. Planning a rigorous assessment of a study’s implementation outcomes, beginning with study recruitment, can further ground study adaptations and facilitate translation into practice. This can be accomplished by proactively and continuously assessing and revising implementation strategies.

## Data Availability

All data produced in the present work are contained in the manuscript.

## ACKNOWLEDGMENTS

The authors are grateful for the hard work of the DETECT-A study team for the successful implementation of a large-scale study, as well as the Geisinger Research departments mentioned in the paper. We also want to thank the 10,006 dedicated women who joined the study and volunteered their time to research.

## Original Protocol IRB #2017-0268

### Contributors

AH, AB, KS, KS, TB, EW, CW, WL were involved with the study design, data collection, analysis, and write-up. AR assisted with the analysis and write-up. All authors contributed to the revisions to the article.

### Funding

This work was supported by Marcus Foundation through Johns Hopkins University.

### Data sharing statement

No additional data are available.

